# Effectiveness of filtering or decontaminating air to reduce or prevent respiratory infections: A systematic review

**DOI:** 10.1101/2023.06.15.23291419

**Authors:** Julii Brainard, Natalia R. Jones, Isabel Catalina Swindells, Elizabeth J. Archer, Anastasia Kolyva, Charlotte Letley, Katharine Pond, Iain R. Lake, Paul R. Hunter

## Abstract

**BACKGROUND:** Installation of technologies to remove or deactivate respiratory pathogens from indoor air is a plausible non-pharmaceutical disease control strategy.

**METHODS:** We undertook a systematic review of observational and experimental studies, published 1970-2022, to synthesise evidence about the effectiveness of suitable indoor air treatment technologies to prevent respiratory or gastrointestinal infections. We searched for data about infection and symptom outcomes for persons who spent minimum 20 hours/week in shared indoor spaces subjected to air treatment strategies hypothesised to change risk of respiratory or gastrointestinal infections or symptoms.

**RESULTS:** Pooled data suggested no net benefits for symptom severity or symptom presence, in absence of confirmed infection. There was weak evidence that air treatment technologies tended to reduce confirmed infections, but these data evinced strong publication bias.

**CONCLUSION:** Although environmental and surface samples are reduced after air treatment by several air treatment strategies, especially germicidal lights and high efficiency particulate air filtration, robust evidence has yet to emerge to confirm that these technologies are effective in real world settings. Data from several relevant randomised trials have yet to report and will be welcome to the evidence base.

## Introduction

Several technologies have been developed or proposed that treat indoor air supplies in a way that might prevent transmission of respiratory infections. Some of the most promising such technologies are safe to operate while people are breathing the same area and moving around in the exposed area. Removing microbes from air is a form of filtration, one example being high efficiency particulate air (HEPA) filtration. As defined by the United States Department of Energy, the HEPA standard is to remove at least 99.97% of aerosols 0.3 micrometers (μm) in diameter (US Department of Energy 2005). Alternatively, rather than remove microbes, an air treatment technology might render microbes incapable of biological replication, and as such, unable of causing infection. Germicidal ultraviolet light (GUVL) in bandwidths safe for chronic human exposure but also able to deactivate viruses, has been proposed as such a way to decontaminate air from pathogens (Narita et al. 2020).

During the Covid-19 pandemic, such air treatment technologies were promoted as a practical mitigation measure in environments where social distancing was difficult to maintain; many governments at local and national level announced support for such technology to be deployed widely, especially in schools (Camfil 2021, Ulmair 2021, Zimmer 2021). These aspirations were hindered by the large cost involved in providing suitable air treatment devices in all proposed settings and uncertainty about exactly which devices might be truly effective (Brandon 2020, Akpan & Jeffrey-Wilensky 2021, Wightwick 2021). Some cluster randomised controlled trials to provide possible supporting evidence were subsequently initiated, using either HEPA or GUVL, in schools (ISRCTN46750688; NCT05016271) or long-term residential care homes (ACTRN12621000567820; NCT05084898; ISRCTN63437172). These trial results are not yet available.

Any proposed novel technology or treatment, such as vaccination or a new drug, needs to go through many stages of development, including rigorous safety testing and real-world experiments, before effectiveness is established and large population treatment is justified. Technologies that may purify/treat air are rapidly evolving and are concurrently at all stages of development. We undertook a systematic review updated with evidence available through late 2022 about effectiveness at the application stage, describing respiratory and/or gastrointestinal infection outcomes in human beings following exposure in real world settings to air purifying/treatment strategies and technologies. We consider a broad range of potential technologies and both observational study designs (cohort or case control) as well as experimental trials. We consider exposures whether the technology is portable or a permanent feature of the setting.

## Methods

We sought studies published in 1970 or later, using Google Scholar, OVID MEDLINE, Scopus, medRxiv, bioRxiv, preprints.org. Grey literature published by December 2022 was also searched; trial registries (NCT, ISRCTN and ACTRN) were searched in June 2022. Details of the search terms and parameters are in the Appendix. Eligible studies could be written in any language in which we had literacy (English, Spanish, Greek, French, Italian) or that we could fully translate into English using Google Translate.

Study design had to be controlled experiments, case-control or cohort studies with concurrent comparison groups. Pre-post comparisons were excluded because changes in other conditions cannot be controlled for (Thiese 2014).

Study titles and abstracts were screened independently by two authors to decide which ones to take to full text review. A third researcher was consulted if disagreements could not be resolved by discussion. Full texts of studies not excluded from title/abstract screening were obtained where possible and reviewed for eligibility. A protocol was registered in association with this review (Prospero CRD42020208109); however, we had substantial protocol deviations due to resource constraints and improved understanding of the relevant literature. Further details on study selection are included in the Appendix.

### Risk of bias (quality) assessment

Quality assessment approach depended on study design. Trials were assessed for risk of bias using the Cochrane risk of bias tool 1.0 (Higgins & Altman 2008), with an additional domain for adherence (low risk of bias if reported to be ≥ 64%). One point was awarded for each domain with low risk of bias, and trials with least risk of bias were deemed to be those studies with scores ≥ 6. The quality checklist used for observational studies (cohort or case-control design) was based on the Newcastle Ottawa Scale (NOS; Wells et al. 2000) with a modification that the comparability domain was a single checklist item, whether the groups were balanced at baseline for age and sex.

### Outcomes

Eligible outcomes related to incidence of respiratory/gastrointestinal infection or compatible symptomatic illness in humans, in the context of whether the cohort had been exposed (or not, concurrently) to an eligible technology that treated, decontaminated or filtered air. Included studies had to report at a minimum, the mean effect value for exposed/control cohorts; studies that collected relevant data but did not report raw outcome data or change from baseline, or that only reported between group differences after adjusting (in their own models) for possible confounders were ineligible.

Preferred outcome was incidence (dichotomous yes/no) of respiratory/gastrointestinal infection by a specific pathogen (such as influenza or norovirus) confirmed by a laboratory method. If laboratory-confirmed infection data were not available, we accepted respiratory symptoms such as: cough, acute breathing difficulty, anosmia, rhinitis, nasal congestion, scores for combined respiratory disease symptoms. Eligible gastrointestinal symptoms were nausea, abdominal cramping, vomiting, or diarrhoea that could not be attributed to non-infectious cause. Symptoms could be expressed a dichotomous or continuous (severity) data. Further description of outcomes are in the Appendix.

### Intervention(s), exposure(s)

Eligible technology was treatment of indoor breathing air deemed suitable for use while humans were present doing routine activities (such as sleeping, working, eating, studying) without specialist protective equipment. Treatment methods that required humans to vacate the space during operation of the technology or chemical application to surfaces, or that required special protective equipment for humans to remain present, were ineligible.

Eligible technology could be radiation, chemical, or mechanical systems that aimed to safely purify the air freely circulating in the indoor environment without simply ventilating (putting old indoor out & bringing new air in). Exemplar technologies and treatment methods are HEPA filters, ionisers, GUVL in safe bandwidths for recurring exposure (Narita et al. 2020), and some types of chemical treatment. Studies that describe disinfection systems that move air to a private space where it may be exposed to chemicals/radiation/physical filter were eligible as long as these systems could operate while persons were present in the environment meant to receive the disinfected air AND the populated spaces that received the disinfected air normally received this disinfected air within two hours of treatment. Two hours was not meant to be a definitive threshold, but rather a maximum reasonable period that still enabled the air processing to be relatively quick.

In absence of contrary information, we assumed that any air conditioning system was likely to include some amount of air filtration as part of routine operation, although we could not know how filtered the air was if not explicitly stated.

### Settings

This technology must have operated in a non-laboratory setting, and must have been designed to potentially be applied to an air space shared by five or more persons. This stipulation about size of population exposed was applied because we wanted to exclude cases of specialist negative pressure rooms, small spaces under laminar flow tents, or other resource-intensive, typically clinical/laboratory environments that are typically intended to create very sterile conditions for a single patient or experimental participant. Outcomes had to be in people. Virions or other pathogens in air had to be removed directly from the air, not observed to be reduced after pathogen removal from surfaces or from standing water in the shared environment. Incidence of microbes on surfaces or in air samples were ineligible outcomes.

We excluded observational studies about workers in a small number (< 12) of different buildings, in the context of ‘sick building syndrome.’ Often these studies considered correlation between respiratory symptoms and presence of air conditioner filters, which were theorised to be clogged with harmful dust or pathogens, and otherwise hindering ventilation. However, other factors that affect air quality, both unobserved and observed, were reported to be highly heterogenous, such as concentration of volatile organic compounds, temperature, humidity, density of staff, types of office equipment and ventilation rates. Our own study was not designed to adequately address this diversity of confounding in clustered cohort studies.

### Intervention: Minimum Exposure

The majority of the intervention group had to be present in the setting where air was disinfected for a mean duration of at least 20 hours a week during the monitoring period (about 12.5% of a person’s lived hours per week). The persons could be present for any reason (such as residence, education, work, receiving inpatient treatment, etc).

### Comparator(s)/control

The comparator group had to simultaneously experience usual ventilation regimes in same or similar settings, so exposed to systems that manage air flow but did not attempt to disinfect air or remove microbes from the air. Simple mechanical ventilation (i.e., expelling indoor air and replacing it with outdoor air) was the ideal comparator exposure.

### Synthesis

We summarise the data narratively and quantitatively. All trials (randomised or not) are grouped for synthesis; all observational study designs are grouped together. Where suitable data were supplied (participant count in each exposure group, event count or mean effect and standard deviation/error for ratio outcomes) in at least 2 studies of same design assessing a specific type of air treatment method and outcome, we carried out random-effects meta-analysis with Review Manager version 5 (RevMan 2014). Studies with results that were too incompletely described to synthesise with other evidence are described narratively.

The diversity of reported respiratory symptoms meant that pooled analysis was often only possible by grouping similar measures. To enable synthesis, outcomes were grouped by similarity under three possible umbrella terms: gastrointestinal; laboratory or clinical diagnosis of respiratory infection and/or pneumonia, bronchitis; other respiratory symptoms. The direction of scales in synthesis forest plots was standardised so that a lower value signified less illness/fewer symptoms. Where one study reported multiple eligible outcomes, we did not count the same participants twice in synthesis. We extracted both continuous and dichotomous outcome from the eligible studies. Further description of the synthesis methods are included in the Appendix.

## Results

Study selection is in Figure 1. From 39,346 initial bibliographic and grey literature hits, we found 32 eligible studies within which 41 outcomes were compared between groups. All included studies were either trials or cohort design (no case-control studies). All outcomes related to respiratory infections or symptoms, except for one study in care homes, which looked for norovirus outbreaks related to air conditioning status. Studies are described in Table 1 by type of outcome, technology, and study design (which is how they were grouped in synthesis). The median year of publication was 2008, with seven studies published after 2013 (in most recent ten years). Six studies were about research undertaken after 2013. Eleven studies took place in North America, 9 in Europe, 12 elsewhere (Canada, Singapore, China, South Korea, Hong Kong, Israel, Australia). Exposure settings were private homes (n=16), offices (n=6), clinical (n=5), childcare providers or schools (n=3) and shared residences (care homes or military barracks, n=2). Technologies were HEPA standard air filtration (n=14), filters as part of air conditioning (not specified as HEPA standard, n=8), GUVL (n=3), Ionisers (n=4), laminar air flow filter and air flow system with or without HEPA standard (n=2), electrostatic cleaner (n=2) and chemical (mugwort leaf smoke, n=1); sometimes multiple air treatment technologies were applied simultaneously. One article was in Chinese; all other articles were written in English. Study designs were controlled trials: 25, cohort: 7. 26 studies provided data suitable for pooling (with participant counts, unadjusted mean effect size, variance indicator such as standard error or deviation on effect size).

**Figure 1.**
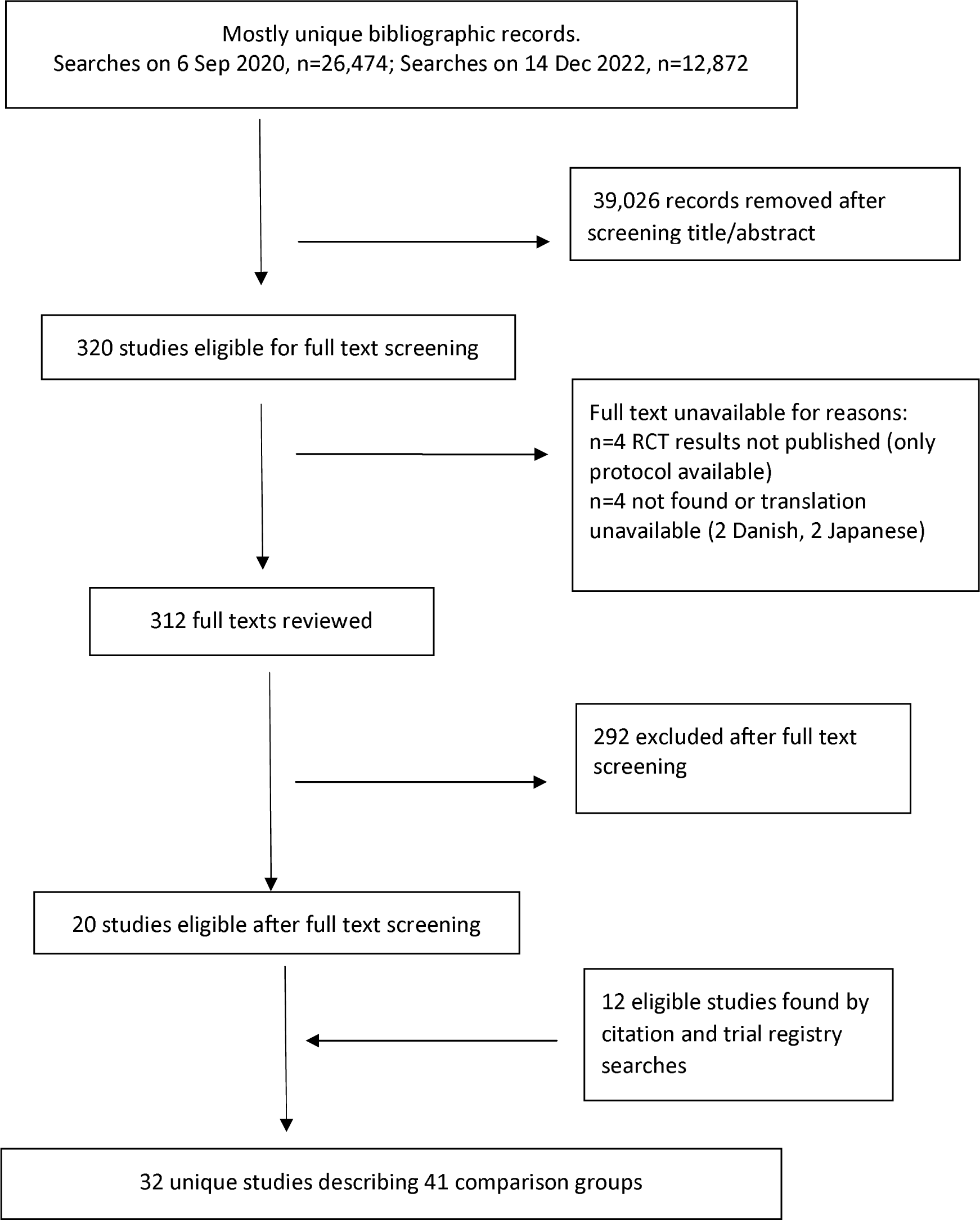
Selection procedure for eligible studies

**Table 1.**
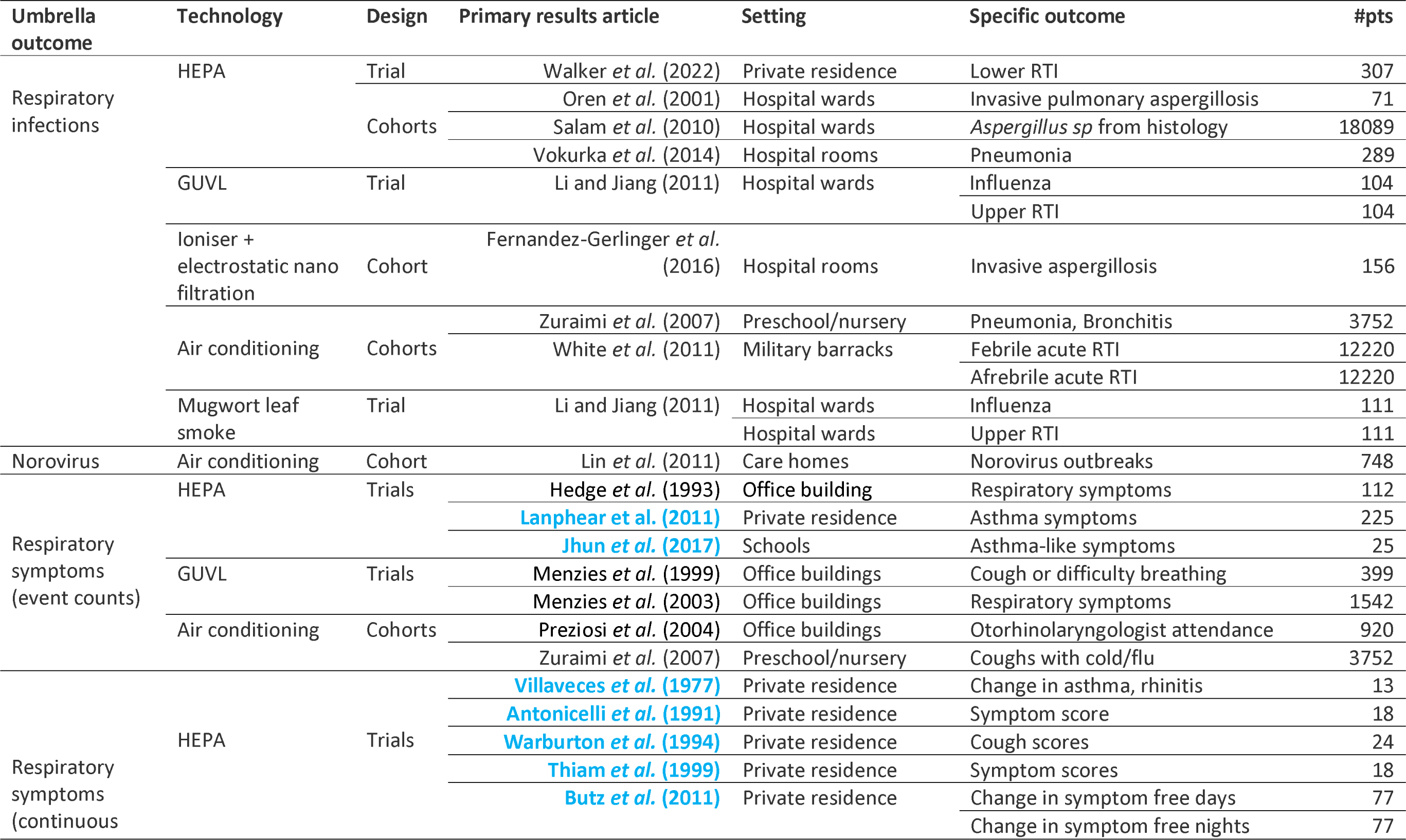
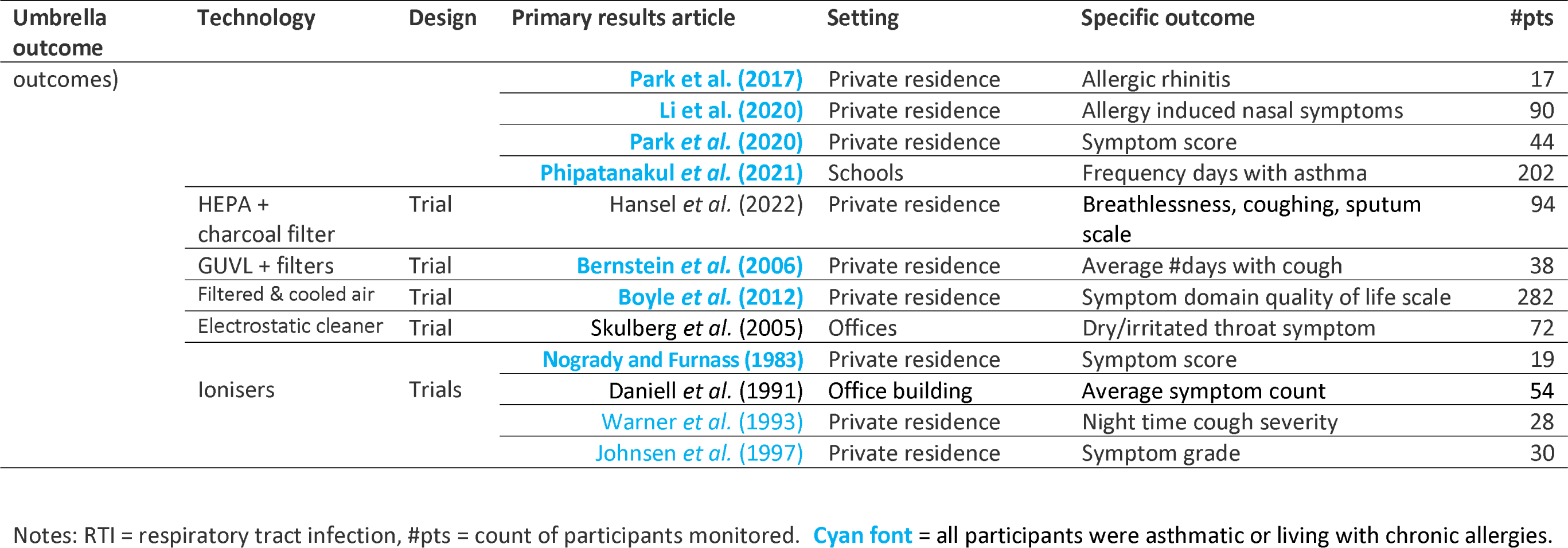
Included studies, technologies, outcomes and participant counts

### Quality Assessment

We used grouped umbrella outcomes as shown in Table 1. Risk of bias assessment is in Table 2 (trials) and Table 3 (cohort studies). Figures 2a-2c show funnel plots for the meta-analyses in Figures 3-5. Figure 2a (pertaining to data shown in Figure 3) suggests strong publication bias (imbalanced distribution of effect sizes; Malički & Marušić 2014) for infection outcomes, but publication bias is not obvious for symptomatic outcomes (funnel plots 2b and 2c, pertaining to data used to construct syntheses in Figures 4-5).

**Figure 2.**
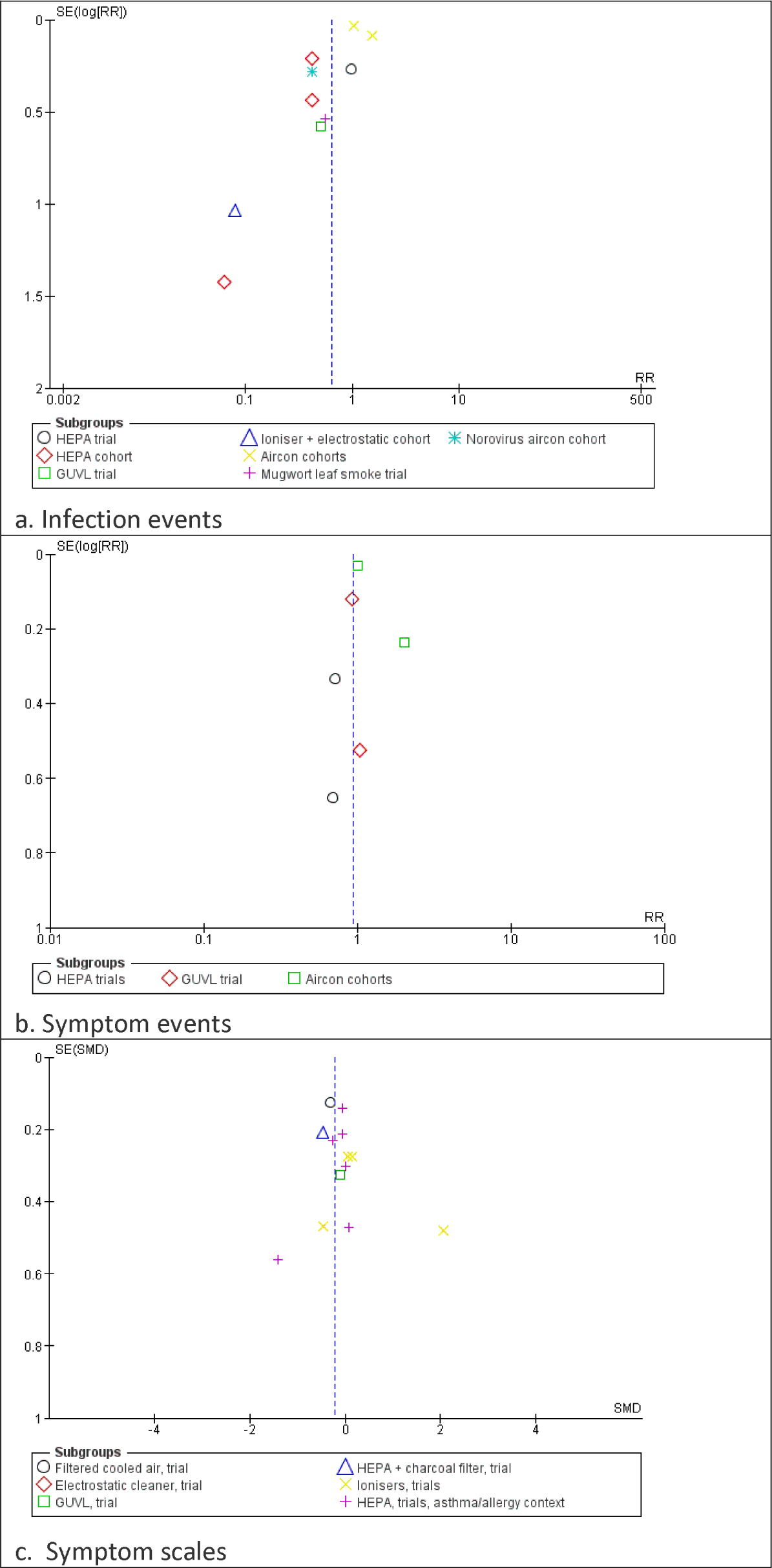
Funnel plots for studies shown with infection events (2a: Figure 3 data), symptom events (2b: Figure 4 data) or symptom scales (2c: Figure 5 data).

**Figure 3.**
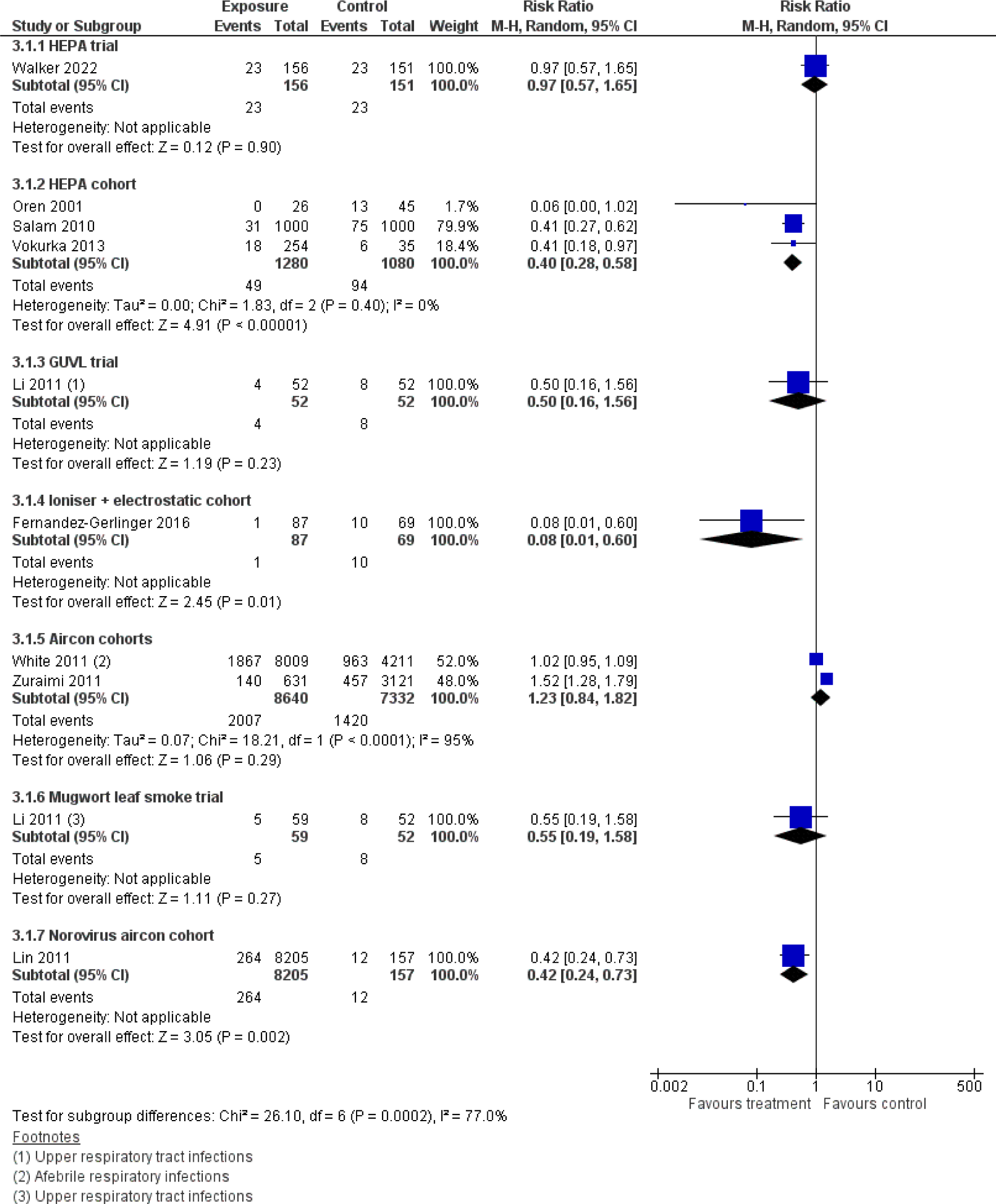
Infection outcomes

**Figure 4.**
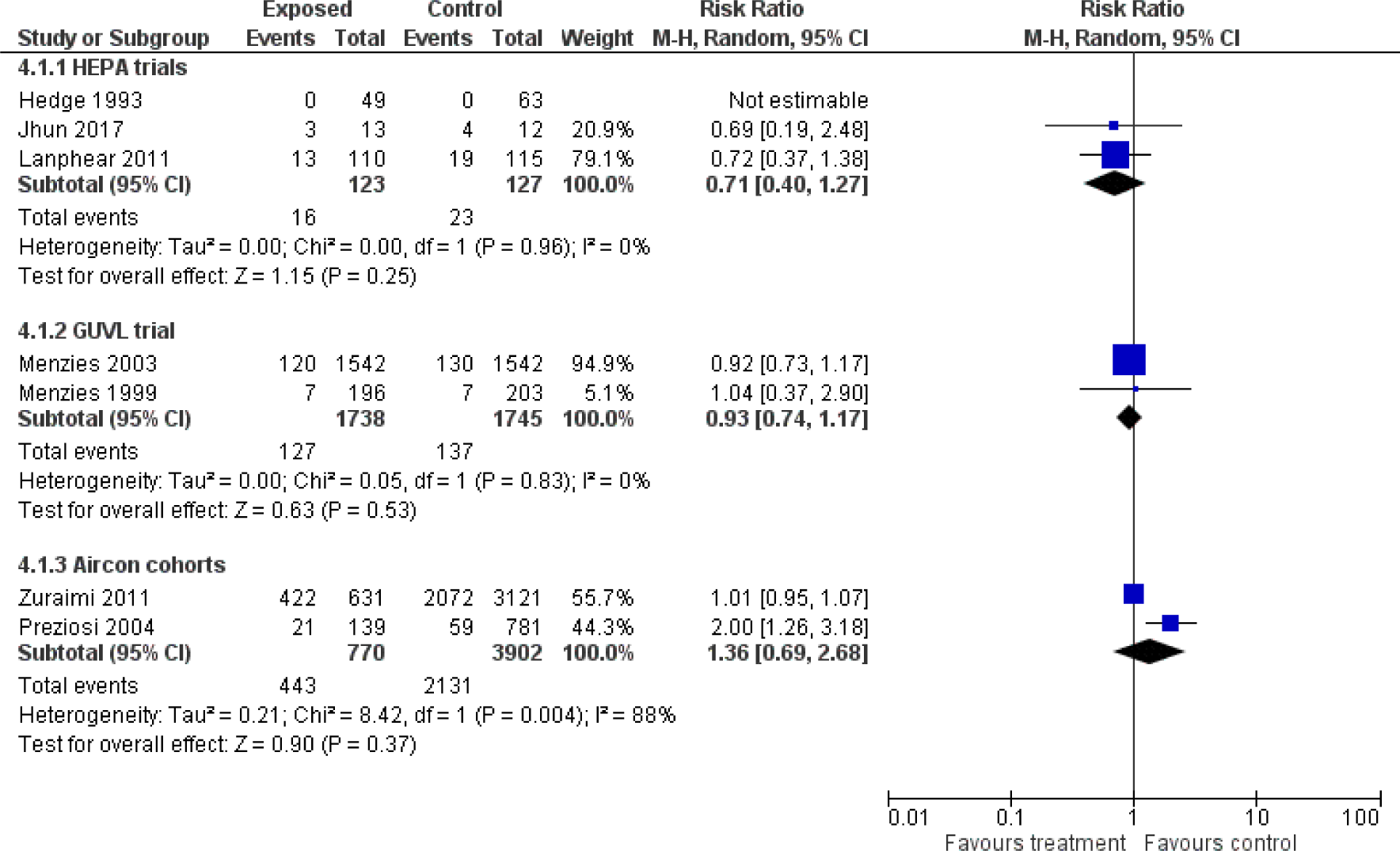
Symptoms as dichotomous outcomes

**Figure 5.**
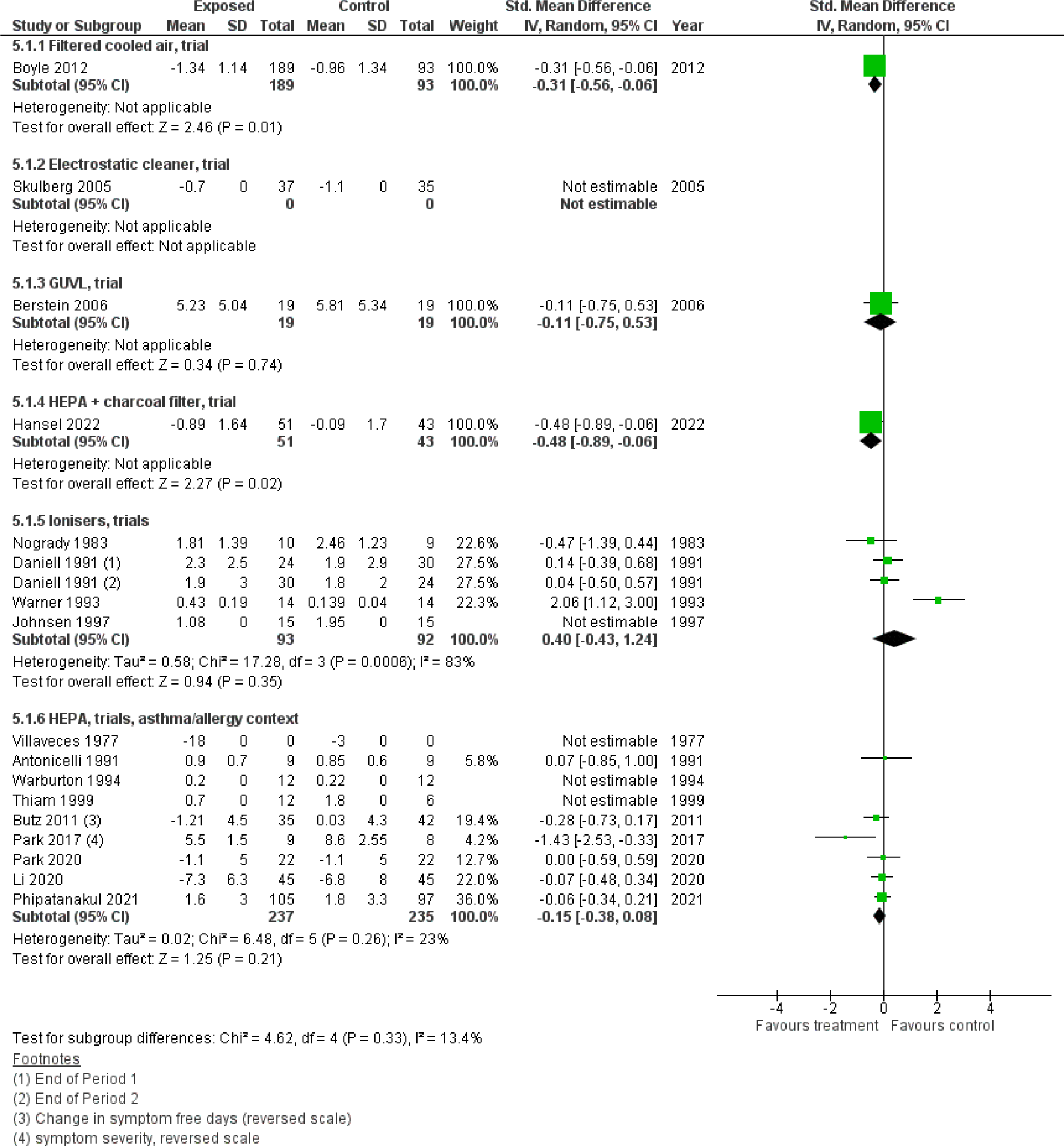
Respiratory severity scores

**Table 2.**
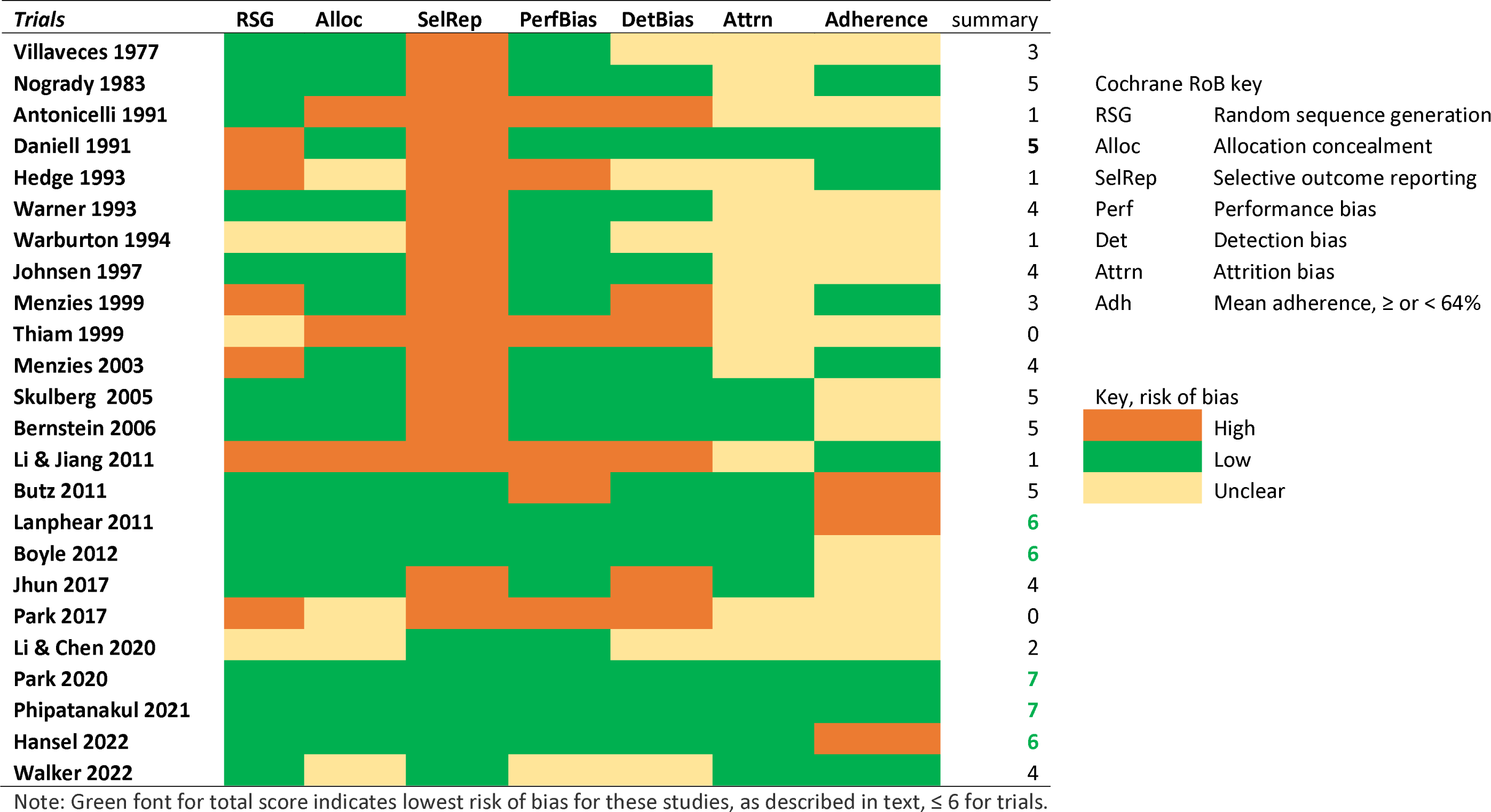
Risk of Bias for controlled trials, Cochane RoB 1.0

**Table 3.**
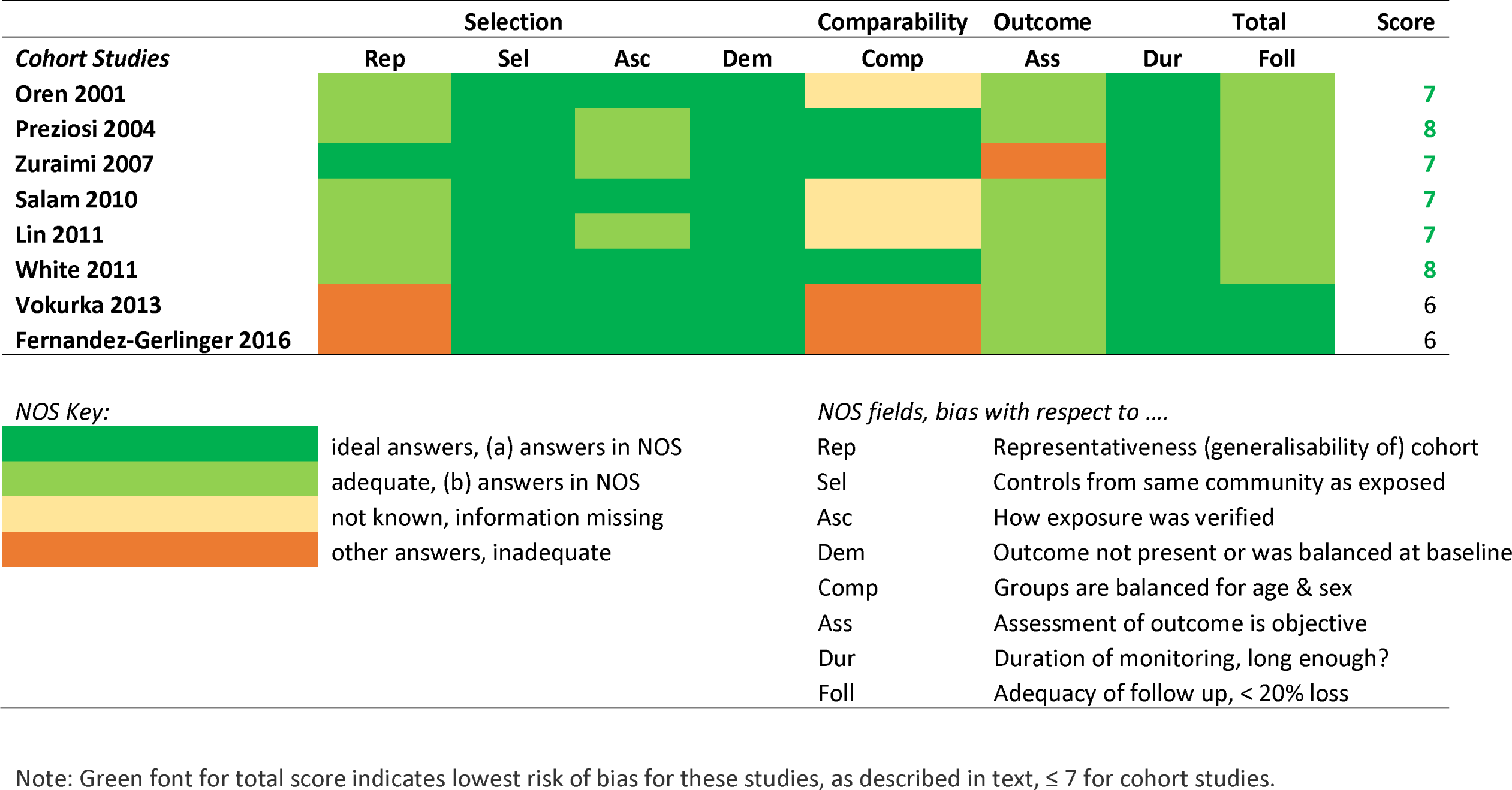
Risk of Bias for observational studies, Newcastle Ottawa Scale

#### Synthesis and outcomes

Figure 3 shows pooled risk ratios for infections as outcomes, with subgroups by umbrella outcome, study design (trial or cohort) and technology. There was a trend towards the treatment groups to have fewer infections. This finding was more consistent for observational studies, especially HEPA cohorts. Confidence in the HEPA cohort comparisons can be boosted because of their low heterogeneity (I2 = 0%); in contrast to the high heterogeneity (95%) in the air conditioning cohort comparison for respiratory infections. Ionisers with electrostatic technology also appeared to have a strong protective effect, however this finding is from only one moderate size study which was not undertaken for a highly generalisable group (care home residents, which groups were unbalanced for sex/age at baseline). No trials had effects that were in favour of air treatment to reduce infection at p < 0.05. We also note (Figure 2a) that there is strong evidence of publication bias in this group. The only gastrointestinal study we found was for norovirus outbreaks. Comparison 3.1.7 found fewer norovirus outbreaks in care homes with air conditioning; however, this result may be interpreted with caution given that only a small percentage of participants lived without air conditioning.

Figure 4 shows pooled data for dichotomous respiratory symptom outcomes. There was no overall trend towards favouring controls or treatment. Heterogeneity was especially high (I2 = 88%) for the air conditioning as treatment method, not dissimilar to the high heterogeneity for air conditioning treatment in Figure 3.

Figure 5 shows respiratory symptomatic severity outcomes expressed as continuous data (scores or scales; higher is worse), using standardised mean differences. Between group effects could not be estimated for some studies because variance data were unavailable in Skulberg et al. 2005, Villaveces et al. 1977, Warburton et al. 1994, Johnsen et al. 1997 and Thiam et al. 1999. Most studies did not find statistically significant evidence to support treatment effect in reducing symptom severity. Combined air treatment (such as cooled and filtered, or HEPA with additional charcoal filtration) seemed to perform better than single technology approaches (e.g., just HEPA or ionisers). Filtered (non HEPA) and cooled air had the best results in terms of reducing symptom severity. The mean effect in Boyle et al. 2012 was –0.31 (95%CI –0.56 to –0.06). One study gave especially strong support in favour of HEPA treatment for asthmatics (Park et al. 2017). Evidence was especially heterogenous for ionisers (I2 = 83%), with the untreated groups tending to have fewer symptoms (pooled SMD 0.40, 95%CI –0.43 to 1.24).

#### Costs and maintenance

Most studies (n=28) made no statement about costs of the technology. Menzies *et al*. (1999) said that GUVL was a “relatively low cost intervention”. (Menzies *et al*. 2003) were more specific, saying that to install GUVL in an office building with 1000 staff would cost circa (USD) $52,000 to install and about $14,000 in annual running costs (electricity and replacement bulbs), resulting in an investment cost of $52 and annual running costs of $14 per employee. With respect to HEPA filtration, Salam *et al*. (2010; device used in private homes) said that two portable HEPA filtration units cost about $900 each with annual running costs circa $500, while (Butz *et al*. 2011; devices used in hospital rooms) said that likely costs were $200-$400 per installed unit. Authors relied on citation of other documents to address sustainability or maintenance issues related to device, operation, although Jhun *et al*. (2017) said that the HEPA device filters only needed to be changed once a year, while Park *et al*. (2017) said that HEPA filters had been changed during the intervention period after 12 weeks.

#### Adverse effects

Most studies (n=21) did not comment on whether adverse effects of the intervention or exposure had been looked for or analysed. Of studies that did discuss possible adverse effects, noise from devices was the most common participant complaint (n= 4). Nogrady and Furnass (1983) reported lack of ozone being detected, related to ioniser use. Li *et al*. (2020) undertook especially systematic data collection for device tolerability, with weekly Likert scale questions about whether the device operation was tolerable. Eye irritation from mugwort smoke was mentioned in Li and Jiang (2011).

## Discussion

Our review has novel findings not previously evaluated using rigorous systematic review methods. A systematic review of literature published by late 2020 Hammond *et al*. (2021) concluded that no existing studies had yet investigated incidence of respiratory infections using portable HEPA filter devices. Our literature search is both updated and applies much wider inclusion search criteria than those applied in Hammond *et al*. (2021) because we included both portable and installed in situ technologies, as well as a much wider types of technologies (GUVL, ionisers, electrostatic cleaners, etc.). Our review also considered a broader range of outcomes, respiratory symptoms (severity scales or incidence) as well as incidence of respiratory infections.

Air treatment technologies that successfully inactivated SARS-CoV-2 in air samples and on surfaces has been widely reported, including studies in community settings (Rodríguez *et al*. 2021, Myers *et al*. 2022, Zhang *et al*. 2022). We did not systematically collect or review environmental outcomes evidence, but agree that much of this research suggests that some technologies can be very effective at reducing microbe presence in environments while people are undertaking routine activities. However, while those environmental sampling results are promising, our synthesis of data to date about symptom and infection outcomes in human beings could not confirm that air purifying/treatment technology is likely to reduce respiratory or gastrointestinal infections. This finding could be, in part, due to a lack of rigorous evidence, a deficit that should be redressed by 2026. Where symptoms or infections seemed to most reduce was in association with combined technology, such as ionisers with electrostatic cleaners, or HEPA standard filters with additional charcoal-based filtration.

Controlled swine farms studies found reduced clinical signs of enzootic pneumonia, atrophic rhinitis and other viral indicators among animals subject to air filtration (HEPA or MERV rating 14 / 16) and resident in the facilities at all times (Lau *et al*. 1996, Dee *et al*. 2012). It is encouraging that air treatment technologies in these types of environments reduced airborne infections. We note that a key difference between a livestock farm and human activities is that most humans are not confined to a single indoor space for weeks or months, with large groups of similarly confined co-residents. Exceptions are prisoners and in general, many care home residents. One Portuguese study found that elderly care home residents in 2014 spent an average 95% of their time indoors at their residential facility (Almeida-Silva *et al*. 2014). In these unusual environments, technologies that try to stop disease transmission by disinfecting air have the greatest chance of success.

Six of our included studies reported on data collected in 2014-2023. However, we found just four reports about experiments at any date (rather than observational study designs) that collected data about infections in humans after air treatment that was meant to deactivate or remove pathogens from indoor air. Lack of rigorous experimental trials is problematic because of the greater biases in cohort (observational) study designs, but even in randomised controlled trials (RCT) study designs, biases introduced by poor randomisation, blinding and allocation concealment may exceed the apparent preventive effects suggested by the cohort studies included in our review (Wood *et al*. 2008, Savović *et al*. 2012). Nevertheless, it is excellent that at least five cluster RCTs were registered since 2020, in four different countries, to evaluate the most recent technologies that may be able to adequately remove/deactivate contagions in shared indoor spaces. These trials will have evaluated both HEPA (n=3) and GUVL (n=2). According to registrations, two school-based trials (ISRCTN46750688; NCT05016271) were scheduled to finish data collection by late 2022; while three experiments in care home settings (NCT05084898; ACTRN12621000567820; ISRCTN63437172) will finish data collection in 2023 and 2024. Unfortunately, we did find evidence of publication bias in the existing evidence base.

We did not expect to find so many studies undertaken in the context of allergenic response or asthma. We felt we should include these studies unless the authors said they had excluded infection as a cause of symptoms (which they did not). We also realised that we needed to include for full text review all studies about respiratory or gastrointestinal outcomes in people where a relevant technology was tested in an eligible setting. Even if our outcomes were not mentioned in the article abstract, these data were sometimes collected and reported in the full report. Reviewing full text of so many articles exceeded our initial resource allocation. We also decided that it was undesirable to confine our review to only dichotomous outcomes as stated in the original protocol. These are among the many reasons for deviating from our original protocol (Prospero CRD42020208109).

Operational costs are not a small concern when a technology is suggested to be rolled out at scale, as happened during the Covid-19 pandemic. The costs of providing these air disinfection solutions are likely to still be prohibitive in many settings (Wightwick 2021, Zimmer 2021). In future, costs are likely to reduce, and may coincide with the time point when the technology is proven to give effective protection against disease transmission. Resource limits are an uncomfortable reality with regard to any medical or public health intervention: data on implementation costs should be included in published evaluations.

There is a distinct lack of studies addressing aerosol or other possible airborne transmission of gastrointestinal infections. Although aerosol transmission is a lesser pathway for gastrointestinal infections, it does happen, notably following project vomiting, for instance, which is often associated with norovirus infection (Makison Booth 2014). Norovirus outbreaks have been associated with air travel in spite of HEPA filtration being routinely fit on nearly all commercial aircraft manufactured in recent decades (Thornley *et al*. 2011). Experiments evaluating effective protection from air treatment systems should consider many common very transmissible pathogens, and in so-doing may establish greater benefits.

Potential adverse effects in earlier experiments were often simply not addressed, not commented on at all. The most common adverse effect reported, which sometimes led to trial withdrawal, was noise. This potential nuisance is likely to be reduced with technological developments.

### Strengths and Limitations

We undertook a very large search in diverse bibliographic sources (engineering, environmental, medical and health sciences), including three trial registries. We looked beyond abstracts to find evidence of outcomes in humans regardless of whether or not the abstract indicated those outcomes had been monitored. We undertook thorough forward and backward citation searches; about a third of our included studies came from citation searches. We searched nine good similar systematic reviews for additional studies to include.

We made inclusion decisions that could have slightly adjusted findings. We excluded studies published before 1970; we are aware of 1940s-1950s studies with both encouraging and equivocal results using GUVL (Reed 2010). We were unable to incorporate results from at least five very modern trials (initiated ≥ 2020) that have yet to report. Contacting original authors for additional information exceeded our resource capacity, although we note that most studies were published before 2010 so it is unlikely that much additional information could have been collected. We excluded articles that did not report primary raw (unadjusted) outcomes. We excluded multifactorial experiments, such as Eggleston *et al*. (2005), which had HEPA filters as well as environmental actions in the only intervention arm. We found many studies that collected symptom outcome data related to air treatment but did not report raw results we could input to group synthesis. For instance, Shao *et al*. (2017) collected data about shortness of breath in participants, but did not report this information. In models adjusted for participant age and gender, Noonan *et al*. (2017; RCT in homes) found no improvement in (asthmatic) symptom severity related to HEPA filtration. In models adjusted for 13 other covariates, Abd Razak *et al*. (2020; cohort study in child care centres) found greater symptom severity related to air conditioning rather than natural ventilation. Gent *et al*. (2022; RCT in homes) found reduced symptomatic illness related to HEPA filtration in homes of asthmatics, after adjusting for measured NO2 concentrations in same environment. These findings suggest that adjustment by many types of confounders may be warranted to find true effect size, a study design that requires access to individual participant data. We did not attempt individual participant meta-analysis, but otherwise note that the lack of apparent consensus from these other studies is similar to our own findings.

We have not attempted to adjust for aspects of any study such as participant vulnerability, participant ages, device air flow rates, person-hours of exposure, adherence to trial protocol or vulnerabilities of target pathogens, any one of which may well affect real world effectiveness. We endeavoured to undertake our synthesis with minimal bias but acknowledge that it is not ideal that our own study did not adhere to a pre-registered protocol.

### Conclusions

Evidence that treating indoor air in public spaces will reliably prevent transmission of respiratory or gastrointestinal diseases remains elusive. Pooled data suggested no net benefits for symptom severity or symptom presence, in absence of confirmed infection. There is weak evidence that air treatment technologies tended to reduce confirmed infections, but these data evince strong publication bias. Although environmental and surface samples are reduced after air treatment by several air treatment strategies, especially germicidal lights and high efficiency particulate air filtration, robust evidence has yet to emerge to confirm that these technologies are effective in real world settings. Data from several relevant randomised trials have yet to report and will be welcome to the evidence base. Where such technology is trialled, costings and adverse events should be reported to contextualise any potential trade offs in public health protection decisions. We recommend that authors publish both raw unadjusted outcome measures as well as results from appropriately adjusted models to facilitate multi-study synthesis.

## Data Availability

All data produced in the present work are contained in the manuscript

## Declarations

### Approval to use the data to undertake the research

Approval was not required because this is secondary analysis of published data.

### Author contributions

PRH conceived of the study. IRL and PRH secured funding. JB co-designed and ran the searches. JB integrated and de-duplicated bibliographic hits. JB, NRJ, ICS, EJA, AK, CL and KP screened titles and abstracts. ICS undertook backward and forward citation searches with confirmation by JB, who also checked references of systematic reviews for additional studies. JB and NRJ screened full text. JB and ICS initially extracted data from full text, confirmed by each other or NRJ. JB and PRH designed the synthesis strategy. JB and NRJ undertook quality assessment. JB wrote the first draft and assembled revisions with comments from all coauthors. All authors have read and approve of the final manuscript.

## Acknowledgements

Thanks for advice from colleagues at Norwich Medical School and Matteo Carpentieri at the School of Mechanical Engineering, University of Surrey.

## SEARCH STRATEGY

**Google Scholar** (first 20 hits only for each combination, including links to patents) respiratory /or/ gastrointestinal /or/ norovirus

and each separately from this list

air disinfection /or/ air cleansing /or/ air filtering/or/ air filtration

and each separately from this list…

germicidal; air purification; laminar; HEPA; far UVC, hydrogen peroxide

**Scopus** (all hits back to 1970)

(TITLE-ABS-key

((*respiratory* OR *coronavirus* OR *influenza* OR *legionella* OR *tuberculosis* OR *common-cold* OR *rhinovirus* OR *norovirus* OR *vomit\** OR *diarr\** OR *gastrointes\** OR *sick\** OR *ill\**))

AND

TITLE-ABS-KEY (*purification* OR *disinfection* OR *filtration* OR *germicidal* OR *laminar-flow* OR *hepa* OR *far-uvc* OR *hydrogen-peroxide*) AND TITLE-ABS-KEY (*trial* OR *rct* OR *experimen\** OR *effect\** OR *case-control* OR *cohort* OR *longitudinal*))

AND

(LIMIT-TO (DOCTYPE, *“ar”*)) AND (LIMIT-TO (SUBJAREA, *“MEDI”*) OR LIMIT-TO (SUBJAREA, *“ENGI”*) OR LIMIT-TO (SUBJAREA, *“ENVI”*))

AND

(LIMIT-TO (LANGUAGE, *“English”*))

**medrxiv, bioRxiv, preprints.org** (used exact phrase on preprints.org, first 75 only on bioRxiv sorted by best match) (in each respository) air-filtration or air-disinfection or germicidal or irradiation or laminar-flow or ultraviolet or far-UVC or HEPA or peroxide

**Trial registries** (ANZCTR, NCT, ISRCTN) were all searched on 10 June 2022 and for updates on same trials on 5 Dec 2022, with these terms: (ultraviolet or filter or hepa) and (respiratory or covid)

## OVID MEDLINE

Database: Ovid MEDLINE(R) and Epub Ahead of Print, In-Process & Other Non-Indexed Citations, Daily and Versions(R) <1946 to 6 Sept 2020<

Search Strategy:

1 exp Air Filters/ (350) 2 exp Filtration/ (36555)

3 2 and 1970:2012.(sa_year). (27640)

4 ((purify or purifi* or disinfect* or filtrat* or sanitiz* or sanitis* or ioniz* or ionis* or filter* or germicid* or laminar-flow* or HEPA or irradiat* or far-UVC) adj5 air*).ti,ab. (7691)

5 1 or 3 or 4 (34802)

6 (respiratory or coronavirus* or influenza* or flu or legionella* or tuberculosis or TB or (common adj3 cold*) or rhinovirus*).ti,ab. (770505)

7 exp Respiratory Tract Infections/ (378001) 8 6 or 7 (979407)

9 control groups/ or double-blind method/ or random allocation/ or single-blind method/ or exp case-control studies/ or exp cohort studies/ (2474810)

10 randomized controlled trial.pt. (512469) 11 controlled clinical trial.pt. (93828)

12 randomized.ab. (491495) 13 placebo.ab. (210767)

14 drug therapy.fs. (2231824) 15 randomly.ab. (340352)

16 trial.ab. (518815)

17 groups.ab. (2089568) 18 randomised.ab. (98135)

19 15 or 12 or 16 or 18 or 11 or 17 or 10 or 13 or 14 (4802048) 20 (animals not (human and animals)).sh. (6660396)

21 9 or 19 (6340283)

22 21 not 20 (5344046)

23 5 and 8 and 22 (245)

24 23 and 1970:2020.(sa_year). (245)

25 (norovirus* or vomit* or diarr* or gastro* or ill* or sick*).ti,ab. (1339654) 26 exp Norovirus/ (4764)

27 25 or 26 (1339940)

28 8 or 27 (2236554)

29 5 and 22 and 28 (365)

## Supplemental Methods

### Study selection

We looked at every full text study that tested an eligible technology in an eligible setting where the abstract said that authors had monitored respiratory or gastrointestinal outcomes, even if the outcomes described in the abstract were not our eligible outcomes. For instance, if a study mentioned results in the abstract only about forced expiratory volume or only about surface swab samples, we checked the full text to see if they authors had also published data within about respiratory symptoms in people (even if not mentioned in the abstract). Data were extracted by a single author and confirmed by others in the research team. Backward and forward citation searches in all included studies was undertaken. We also checked for eligible studies in nine systematic reviews with similar research questions (Lee *et al*. 2005, Eckmanns *et al*. 2006, Blackhall *et al*. 2012, Luongo *et al*. 2016, Cheek *et al*. 2021, Park *et al*. 2021, Bowles *et al*. 2022, Thornton *et al*. 2022). Quality assessment was undertaken by a single author. We did not contact primary study authors for additional information.

### Risk of bias (quality) assessment

Criteria used to assign risk of bias category for each domain are noted in result tables. One point was awarded for each domain with an assessment of low risk of bias. This meant that the scoring for our version of NOS was maximum 8; maximum quality score for trials in our quality assessment was 7 (higher score meant lower risk of bias).

### Outcomes

Study participants with a chronic illness that might present with same symptoms as our outcomes, such as Crohn’s disease or asthma, were included unless infection had been excluded as possible cause. If a study reported compound derived outcome scores, such as a combined score for asthma management, we only considered this compound score to be an eligible outcome if eligible symptoms were dominant in the score derivation. For instance, an asthma management score that had 4 domains of which “night time coughing” was only one domain and the other three domains were laboratory rather than symptomatic measures such as forced expiratory volume, peak expiratory flow and inhaler use frequency: this composite outcome would not be an eligible outcome in our review.

Where one study had multiple eligible symptom outcomes (such as cough and fever and nasal congestion), we preferred them for pooled analysis in this order (most to least preferred): cough, breathing difficulties, nasal congestion, rhinitis/sneezing, throat symptoms, fever, other respiratory symptoms or combined respiratory symptom scores. Preferred gastrointestinal symptoms for synthesis in descending order were: vomiting, nausea, diarrhea, abdominal cramps.

When a study had multiple measurements of an outcome at different moments during the monitoring period, we used cumulative results at the last recorded follow-up within 28 days of latest exposure. Where more than one type of outcome was available (e.g., symptomatic scores and cases or lab-confirmed cases) we extracted both and collated them separately.

We also extracted data about adverse outcomes and comments about costs or maintenance considerations associated with the air treatment technology.

### Synthesis

Dichotomous (yes/no) data were about infection and/or incidence of eligible symptoms. Effectiveness for dichotomous outcomes was expressed as risk ratios on a log scale with 95% confidence intervals. Examples of continuous data were severity scores, frequency or change over baseline. For continuous data, we assessed effectiveness using standardised mean differences with 95% confidence intervals.

Where a study provided multiple eligible outcomes (eg. White *et al*. 2011 reported counts of cases that were either febrile or afebrile respiratory tract infections) we used only one outcome in synthesis, to avoid double counting. We chose in synthesis the outcome with larger absolute values: e.g., there were higher case counts (both arms) for afebrile illness in White *et al*. (2011).

